# Pain Expectations, Experiences and Coping Strategies Used By Post-operative Patients: A Descriptive Phenomelogical Study

**DOI:** 10.1101/2024.02.02.24302133

**Authors:** Richard Sakyi, Edward Appiah Boateng, Abigail Kusi-Amponsah Diji, Kenneth Adjei Afful, Vincent Afriyie Nimoh, Philomena Asakeboba Ajanaba, Mabel Dorothy Adjei, Felix Apiribu, Veronica Millicent Dzomeku

## Abstract

**Objectives:** Post-operative pain is still an unresolved problem worldwide including limited-resource countries such as Ghana. Earlier studies have mainly focused on post-operative pain experiences of patient with little attention on their pain expectations and coping strategies, however exploring these other areas can help to better manage post-operative pain. The current study sought to explore pain expectations, post-operative pain experiences, and coping strategies used by adult surgical patients.

**Methods:** A descriptive phenomenological design approach was used to study nine purposively sampled surgical patients who were receiving care at a regional hospital in Ghana. Participants were individually interviewed before and during the post-operative period to share their opinions on their pain expectations, postoperative pain experiences and coping strategies. The semi-structured individual interviews were audio-recorded, transcribed verbatim and content analyzed to generate themes which described participants’ accounts.

**Results:** Participants comprised of six females and three males; their ages ranged from 24 to 40 years who underwent major surgeries. Three main themes were derived from this study, diverse pain expectations and experiences, post-operative pain effects and post-operative pain coping strategies. The study revealed that participants have different pain expectations and experiences, surgical pain affected their activities of daily living and emotions. Participants coped with the postoperative pain by using personal strategies and seeking support from nurses.

**Conclusion:** Pain expectation of surgical patients affects their post-operative pain experiences. Surgical patients use coping strategies in their post-operative pain management. More needs to be done in reducing surgical patients’ experience of Post-operative pain.

## INTRODUCTION

Post-operative pain is a predictable effect of the injury resulting from surgery and can also be considered as an adaptive response which aids healing by limiting the movements and other behaviors that can cause additional tissue damage(1,2).Post-operative pain can however result in several complications including deep vein thrombosis, pneumonia, delay in healing and discharge and the patient’s inability to get involved in rehabilitation programs, in addition to this is the fact that some patients after various surgical interventions develop chronic pain, (3,4,5).

All patients undergoing surgical procedures expect to experience a certain degree of pain, for this reason post-surgical pain has become one of the greatest concerns for surgical patients (6).Expected higher post-operative pain by patients pre-operatively has been seen to be associated with the experience of stronger post-surgical pain intensity and severity (7). Moreover the experience of post-operative pain by patients is believed to be a worldwide problem(8).Notwithstanding the improvement in pain management interventions, many surgical patients still experience acute post-operative pain(9), in developing countries however the prevalence of postoperative pain is noted to be high(10). This brings to bear the need to do more in order to further decrease the burden of post-operative pain experience by patients.

Patients’ own pain coping strategies is believed to be one patient-centered approach that has been seen to help minimize the pain intensity in post-major surgery as when these coping strategies are effective, they help to produce permanent adaptation (11).Therefore, understanding coping is critical to optimizing healthy development and thus providing the needed support that patients may need to better manage their post-operative pain(12).

One of the goals of postoperative pain management is to ensure maximum function and comfort, this can only be achieved when patients become the center of the care by being actively involved in all decisions and interventions affecting his or her care (13). This is critical in pain management because pain expression is individualized and subjective (14). Hence the need for looking into patients coping strategies.

A review of the literature points to the fact that some work has been done on post-operative pain in other areas in Ghana many of whom centered on nurses and in some cases patients: There is available literature on assessment and management of post-operative pain among nurses in Korle-Bu teaching hospital Accra (15). An ethnographic exploration of postoperative pain experiences among Ghanaian surgical patients in a tertiary health facility and a regional hospital in Accra Ghana have also been done (16). Other available studies have also looked into the knowledge, attitudes, and practices of post-operative pain management by nurses in selected district hospitals in Ghana (17).as well as patient’s experiences with post-operative pain in Komfo Anokye teaching hospital, Kumasi, Ghana (18).

Regarding pain expectations, experiences, and coping strategies used by patients with post-operative pain there is a scarcity of information in Ghana. The findings from this research therefore will provide clinicians and for that matter stakeholders in post-operative pain management further information on the situation of surgical patients’ post-operative pain expectations and their experience of post-operative pain as well as their coping strategies to help improve post-operative pain management.

## Materials and Methods

### Study Site

The study was carried out in the surgical wards of a regional hospital in Ghana. The hospital has an array of medical services including, OPD, 24-hour emergency care, pediatric and neonatal care, obstetrical and gynecological care, general medical services, child and reproductive health care, special clinics (eye, ear-nose-throat, dental, mental health), mortuary services, and theatre /surgical care. It has 330 bed-capacity and conducted about 1083 major surgeries in the year 2020. The hospital currently has 4 surgical wards (male and female surgical wards, orthopedic ward and the gynecological ward).Surgeries which are manage at the male and female medical wards are general surgeries which includes appendectomies,heniorrhaphies,fistulectomies, mastectomies,thyredectomies and colostomies, surgeries like hysterectomies, salpingectomies are managed in the gynecological ward whiles orthopedic surgeries are managed at the orthopedic ward. In total the staff strength of the surgical wards is one hundred and seven nine (179). They run three shifts with each shift having about 5 staffs.

### Research Design

A descriptive phenomenological approach was considered appropriate for the current study as we sought to explore and describe the lived experiences of the participants regarding postoperative pain (19,20).

### Sampling and Sampled Participants

The study involved post-operative patients who went through major surgeries (these are surgeries involving the use of general or spinal anesthesia). A purposive sampling technique was used to sample nine (9) adult patients from the surgical wards of the hospital. Participants were recruited the day before their surgeries. Persons who could neither speak English nor Twi were excluded as well as persons who could not speak audibly and unconscious patients.

### Data Collection Approach

A semi-structured interview guide was used for data collection and recorded with Audio-tape recorder. Components of the interview guide included participant’s demographic characteristics, pain expectations their pain experience and the post-operative pain coping strategies. The interview guide helped to direct the interview process. The questions were formulated taking into account the specific objectives of the study. Probing questions were formulated to elicit deep explanations and understanding of concepts.

The interview guide was pre-tested by using four participants from the target population. The patients who were used for the pretesting were all discharged before the main interviews were conducted. This pre-test was effective in bringing out several ways in which the interview guide could be improved both through the feedback and the experience of actually doing the interview leading to modification of the interview guide. (21).

Most participants were visited on the day before the surgery, as part of the pre-operative preparation to establish rapport with the patients. The study was thoroughly explained to patients and families available. The interviews was conducted in two sessions, before the surgery, patients were interviewed on their pain expectations and subsequently within 48 to 72 hours post-operatively when they have fully recovered from anesthetic influences, and were able to communicate well. However in some cases participants were could not be interviewed preoperatively in such instances they were recruited and interviewed after the surgery. The interviews were conducted in English and Twi which are languages the participants could express themselves well and that the researchers could also understand well. The interviews were conducted at an agreed time and date and mostly at the bed side of participants which were conducive for them when there were also no interruptions from others. It was carried out in the form of a conversation with probing questions following responses in order to guide the conversations towards the objectives of the study. The interviews for each participant lasted between fifteen (15) to thirty (30) munities for both sections. With the full consent of participants, an audio tape recording of the interviews was done. The researchers also wrote down field notes and conducted observations. Participants were also provided with enough time when needed for further explanations, medical records which applied to the study were also collected from participants’ records.

### Data Analysis

The recorded data were transcribed and analyzed using content analysis deductively. The data collection and analysis was done concurrently until saturation that is when no new information was coming out of the interviews. After each interview, the researchers familiarized themselves with the data by transcribing it verbatim after listening to the interview on several occasions, and the transcript was then compared with the audio by listening to the audio and reading through the transcript. (22,23).Two (2) of the interviews were done in English whiles the rest seven (7) were done in Twi. The interviews that were done in Twi were translated and transcribe into English by the researchers after which the transcripts were verified with participants in Twi to ensure that it portrays exactly what they meant. The data was transcribed using Microsoft and analyzed deductively by generating codes categories, and themes taken into consideration the objectives of the studies. From these themes, the researchers developed a textual description of what the participants had experienced. (24).

### Rigour

Qualitative research should be able to meet the following criteria: credibility, transferability, dependability, and confirmability(25).

To ensure credibility the questions were reframed in different ways during the interview to be sure that the answer truly reflects what the respondent’s intentions are, member checking was also done where information obtained was crosschecked from respondents to be sure that the results are true picture of the information they gave to the researchers. A field journal was also kept to keep track of all activities during the research process. To ensure transferability, a thick description of the research context has also been provided. To ensure dependability a thick description of the research process was done, an audit was done where the research process was audited by two experienced nursing researchers. To ensure confirmability a reflective journal was kept and two experienced researchers also audited the research process.

### Ethical Consideration

Approval for this study was obtained from the Committee on Human Research, Publications, and Ethics (CHRPE) of Kwame Nkrumah University of Science and Technology Ghana with reference number CHRPE/AP/259/21. All participants were given the needed explanations and a written consent obtained from them before collecting information from them, all their questions were answered and they were informed of the fact that they are at liberty to opt-out of the study at any point in time and that their names will not be linked to any information they provide.

## RESULTS

### Demographic Characteristics of the Participants

Nine (9) participants were interviewed for this study comprising of six (6) females and three males. Participants’ ethnic backgrounds included: Ewe (2), Gruma (2), Bono (3), Fante (1), and Asante (1). Participants’ age ranged from 24 to 40 years. Type of surgery done included Fistulectomy (1), appendectomy (2), salpingectomy (1), hernia repair (1), and hysterectomy (4).

**Table 1:** Demographic Characteristics of Respondents.

| Participants | Sex | Ethnicity | Type of surgery |
| --- | --- | --- | --- |
| P1 | M | Ewe | Fistulectomy |
| P2 | M | Gruma | Appendectomy |
| P3 | F | Bono | Hysterectomy |
| P4 | F | Fante | Hernia repair |
| P5 | F | Asante | Salpingectomy |
| P6 | F | Bono | Total abdominal Hysterectomy |
| P7 | F | Bono | Hysterectomy |
| P8 | F | Ewe | Hysterectomy |
| P9 | M | Gruma | Appendectomy |

(Source: field data 2021)

### Organization of themes

The table below shows the details of the themes and subthemes that emerged from the data, in all three (3) major themes and eight (8) subthemes emerged from the data.

**Table 2:** Major Themes and Subthemes.

| **THEMES** | **SUBTHEMES** |
| --- | --- |
| **Diverse pain expectations and experiences** | • Expected post-operative pain intensity<br>• Experienced post-operative pain intensity<br>• Sources of information for expected pain<br>• Pain aggravating factors |
| **Effects of post-operative pain** | • Expected post-operative pain effects on patients<br>• Experienced post –operative pain effect |
| **Coping strategies** | • Personal strategies<br>• Support from nurses. |

(Source: field data 2021)

### Diverse Expectations and Experiences

This describes the different expectations participants express with regards to their pain expectations and experience, four(4) subthemes emerged from this major theme which were expected postoperative pain intensity, experienced postoperative pain, sources of information for expected pain and pain aggravating factors they expected pains of varied intensities and actually experienced pains of varied intensities.

### Expected postoperative pain intensity

Participants were expecting different postoperative pain intensities preoperatively. They expected their post-operative pain to be severe, moderate, or mild. Below are some of what participants said:

> *“I know it will be very painful, yes so far as there is going to be a cut even you if you have an injury you will feel the pain so equally surgery you will feel the pain but the surgery pain will be higher than normal injury”*…….**P1**
>
> *“Oh after surgery you will feel pain but during the procedure, because of the anesthesia they give you will not feel pain but I was expecting a moderate pain.”*….. **P**
>
> *“Oh, I am expecting to feel the pain I am thinking that after surgery definitely, I will experience some slight pain.”*……… **P2**

### Experienced Post-Operative Pain Intensity

Participants also described their post-operative pain intensity, it was reported that the pain was severe, moderate, and mild. Participants shared their experiences:

> *“After the surgery when the effects of the anesthesia were wearing off and I return to normal I started feeling severe pain ”…… **P2***
>
> *“The pain was moderate”*………**P5**
>
> *“I did not feel much pain though but intermittently I will feel some slight pain in my lower abdomen”*……. **P8**

### Sources of information for expected pain

Participant’s pain expectations were based on sources which included personal experiences, information received from family members or friends who have gone through similar surgeries as well as their personal intuition. Participants shared their experiences:

> ***“****For me because there is going to be a cut definitely I knew that there is going to be pain”*
>
> ***…….P1***
>
> *“A friend of my did a similar surgery and it was so painful she was even pregnant and also had the fibroid. She was one of my co-workers I visited her while she was in the hospital and it was not easy for her, so when I decide to have this surgery too I discussed it with her and she narrated to me what she went through hmm so I have all this in mine’’…….**P3.***
>
> *‘Because I have down some surgery before an err as I said earlier when you are going in for the procedure during the procedure you will not feel anything because of the anesthesia they will give you but afterwards when the anesthesia finish working you will start feeling the pain so base on the first experience” **P5***.

### Pain Aggravating Factors

Participants also described the factors that aggravated their post-operative pain, for some lying down increases their pain while in the case of others change in position increases their pain, however in the case of others getting out of bed suddenly aggravates their pain.

Participants shared their experiences:

> *..”Right now if I want to stand up I have to take my time if I get up instantly fast then the pain will come”*… **P4**
>
> *….” Sometimes when you are lying and you change position too then you feel the pain more”*…….**P5**

### Effects of post-operative pain

This represented participations expectations and experiences with regards to how the post-operative pain will affect their activities of daily living and how it actually affected their activities of daily living postoperatively. It emerged from the study that they actually experience what they were expecting.

### Expected post-operative pain effects on participants

Participants reported on their expected effects of the post-operative pain. Most participants expected the post-operative pain to affect their activities of daily living which included bathing, brushing of teeth, walking, and lifting. Participants shared some of their expectations:

> *”Yes I know after the surgery I can’t be moving up and down as expected”.***P1**
>
> *“I cannot do things like bathing and walking as I use to be because of the pain, even I might not be able to brush my teeth without assistance”.* **P7**

### Experienced Pain effects

Participants reported on how they were affected by the post-operative pain. The pain affected how they were able to perform their activities of daily living as most could not bath themselves unassisted, and others have difficulty in getting out of bed and moving about. For some, they could not even sleep because of the pain. Participants shared their experiences:

> *… Yes you know because of this pain hmmm I could not do things like bathing, lifting, sleeping, it was not very easy for me at all people have to even assist me in bathing and the others*…. **P3**
>
> *.” mmm, because of the pain when I want to stand it was difficult, I could not do many things oo even I could not bath myself they have to come and clean us”*…. **P7**

### Post-Operative Pain Coping Strategies

This theme describes the strategies participants employed in coping with their postoperative pain. Two (2) subthemes emerged for this theme: personal post-operative pain coping strategies and support from nurses.

### Personal Post-Operative Pain Coping Strategies

Participants reported the personal strategies that were used to cope with their postoperative pain. The study reveals that participants use position changes to cope with their postoperative pain. Participants shared their experiences:

> *“Yes I was in pain I knew that for the pain I should be able to endure it so I tried to endure it, I turn myself in bed, so when I turn to a side the pain reduces small but when I turn more the pain returns then I will turn back”* ……. **P2**
>
> *“Oh for me when I walk around then the pain subsides and when I lie down the pain increases so I usually walk around”.* P3

In addition to the position changes, participants also use other strategies like praying to God, endurance, supporting the site of pain with hand, relaxation, and crying:

> *“Hmm when am lying down and am trying to change my position and I feel the pain I relax, and when am getting up too and I feel the pain I support the area with my hand, so when I was experiencing the pain then I will lie down and be wailing, and sometimes I will support the area with my hand and was also praying in my heard for God to intervene for me to be relieved of the pain.”* **P5.**
>
> *“Hmm you cannot do anything about it when it got to sometimes you have not to plan to cry but you will see yourself crying, but I was also turning myself small when I turn left then I will turn right. I was also murmured”.* **P6.**

### Support from Nurses

Participants also reported to nurses for assistance when coping with their pain, nurses offered support by administration of pain medications, as well as the use of words of encouragement in helping patients cope with their pain. Participants shared when and how this was done:

> *“……. I reported to the nurses and they gave me another shot injection then it help me, yes the nurses also help they will just come to you and say is painful you have to take your time everything will find they will just say nice things to you to calm you down, you to forget your pain”. **P4***
>
> *“Sometimes the nurses will also give you medication to stop the pain. And then they will also be charting with you so that your mind will not be in the pain and some will come and encourage you others too, some of the nurses have gone through the same surgery so they will use their experience to encourage you”. **P5***

## DISCUSSION

The current study sought to explore pain expectations, post-operative pain experiences, and coping strategies used by adult surgical patients. Participants experienced diverse expectations and experiences. Pre-operatively the study revealed that participants expected severe, moderate and mild pain postoperatively. The findings from the current study are consistent with a study by (26) who posited in their study that many of their participants when questioned pre-operatively expected to develop moderate to severe acute pain post-operatively.The expectations in the current study were mainly as a result of some negative information about post-operative pain received from clinicians or others as well as their personal experiences, this can be collaborated by the fact that increased negative pain expectations can be as a result of negative pre-operative information about post-operative pain received from medical professionals(27).Preoperative communication should therefore provide positive information about pain and its management as this can reduce fear and distress and hence set more positive and realistic expectations (28). Clinicians should therefore be aware that improving communication between the health care providers and the patients can go a long way to improve patients’ pain management outcomes (29).

Participants in current study experienced pain of varied intensities (moderate, severe and mild pains) post-operatively. Post-operative pain is mostly the result of physical injury from the surgical incision (1). However several factors including the type of surgery and the pain expectations are associated with the intensity of post-operative pain experienced (7,30). In this current study, participants went through different types of surgeries and also had mild, moderate to severe pain expectations. The findings from the current study are consistent with other studies (16,31) which have also reported that post-operative patients experience pain of different intensities. The agreement of this study with literature reinforces the fact that management of post-operative pain continues to be inadequate (8), clinicians and policymakers should therefore look at other innovative strategies in managing post-operative pain. The fact that expected post-operative pain has been seen to pose a greater risk to actual post-operative pain experience can also aid clinicians and health care practitioners to identify patients at greater risk for post-operative pain for more intensive post-operative pain management (32).

Findings from this study revealed that the post-operative pain affected how participants were able to perform their activities of daily living as most could not bath themselves unassisted, others also had difficulty in getting out of bed and moving about, the post-operative pain also interfered with patients’ sleep and affected them emotionally which led to some of them crying. This findings is consistent with other studies which have also concluded that post-operative pains interferes with patient’s physical activities including activities in bed (turning, sitting up and repositioning) and activities out of bed like walking, sitting in a chair and standing(33,34).as well as their sleep and emotions(35). The consistency of current research with literature can be attributed to the fact that movement restrictions are adaptive responses that patients use to facilitate recuperation and prevent further potential tissue trauma (1, 2). These effects of post-operative pain cannot be underestimated as it has also been reported that immobility can result in the patient developing deep vein thrombosis as well as other complications like pulmonary atelectasis and some psychological disorders (36). These can delay the patients at the hospital and increase the workload in health facilities with its economic consequences. Policymakers and clinicians should therefore see the need for comprehensive pain management interventions to return the patients to his or her normal function and quality (18).

Participants used personal coping strategies and seeking support from nurses to cope with their pain this is consistent with a study by (12) who found that coping strategies can include personal strategies and professional support.

Findings from this study indicates that participants use position changes like turning in bed, stretching, walking and sitting as well as lying down to cope with their postoperative pain, in addition to this they also resorted to supporting the site of pain with their hand. The different position changes use by participants points to the fact that people use different strategies in dealing with similar situations. The findings from the current study are consistent with others studies (37, 10).who asserted that patient’s uses position changes to relieve their pain.

Findings from the current study also indicated that participants, used other strategies like praying, endurance, relaxation and crying to cope with their pain. Some believed that the pain they were experiencing is normal and something that is usually associated with the surgery hence they just have to endure the pain. For some participants’ movements aggravate their pain hence they will prefer relaxing, the findings from this study are consistent with others studies which has concluded that praying (38), relaxation and social withdrawal (12) are coping strategies use by patients, experiencing pain to add to this is the fact that emotion focus coping strategies are more effective in minimizing post-surgical pain (11). It have been seen that strategies like expressing emotions and reassuring thoughts are more related to more perceived control and in turn better psychological well-being. (39).These strategies can be attributed to the fact that positive beliefs and spiritual support can be intended as a basis for hope and hence can justify ones coping efforts in the most adverse circumstances. (40).

These strategies imply that participants attempted to take control for their pain management. This implies that appropriate coping strategies can therefore provide patients with the ability to adjust and be ready to tackle the stressor which may be in the form of pain, damage to body tissue or decrease mobility(11). Clinicians should therefore appreciate the fact that certain coping strategies can help augment the management of postoperative pain as this may help in counseling and shaping personalized pain management to help improve patient recovery(41).

Participants also reported to nurses for assistance in coping with their pain, nurses offered support by administration of pain medications, engaging them in conversations and activities to divert their minds of the pain as well as the use of words of encouragement in helping patients cope with their pain. At the forefront of postoperative pain management are nurses as they play a significant role in the assessment, implementation and evaluation of pain management interventions (42,28) nurses spend 24 hours with the patient, it is therefore not surprising they are the main source of support to participants in coping with their post-operative pain. The fact that patients report their postoperative pain to nurses for support in coping with their post-operative pain in this study is in line with a study conducted by (43) in which it was seen that post-operative patients felt nurses were available and hence sort for help from them in managing their postoperative pain. Seeking professional support in coping with distress is considered a healthy, coping strategy (12). Nurses and for that matter health professionals should therefore be equipped with the needed skills in pain management to provide the needed support to patients in coping with their postoperative pain.

## CONCLUSIONS

This study has concluded that surgical patient’s pre-operative pain expectations have an effect on their actual pain experience after surgery. Clinicians should ensure surgical patients receive positive information about postoperative pain pre-operatively. Patients who expect to experience severe post-operative pain should be targeted for comprehensive pain management. We have further reaffirm the fact that post-operative pain continues to be a problem for surgical patients. Surgical patients rely mainly on internal strategies and external support in coping with their post-operative pain. Nurses are surgical patient’s main source of support in dealing with their post-operative pain. Patients own coping strategies should be incorporated into pain management interventions. Future studies may consider surgical expectations of patients and its impact on their pain outcomes. The effectiveness of pain coping strategies in post-operative pain management can also be explored as well as nurses’ experiences and perceptions on patients pain coping strategies.

### Limitations of the Study

Due to the method that was used for the study. The findings in this study is limited to only this particular setting and hence cannot be generalised. However the processes used in the study have been described in detail in order to make transferability in other settings possible.

## Data Availability

All relevant data are within the manuscript and its Supporting Information files

## Acknowledgement

We thank the patients who gave us their time and the needed information for this work.

**Competing interest**: the authors declare that no competing interest exist.

**Funding**: the authors received no financial support for the conduct of this research.

## AUTHOR CONTRIBUTION

RS, VMD, AKAD, KAA, conceptualized and designed the study.RS, VMD AKAD, KAA, FA and PAA provided methodological insights. RS, VMD, VAN,AKAD, KAA, and MDA coordinated data collection.RS, VMD, MDA, KAA,VAN, EAB,PAA and AKAD carried out the initial analysis and initial manuscript. The results and its intellectual content was discussed and critically reviewed by all authors. The manuscript was critically reviewed and revised by: RS, VMD, AKAD, KAA,VAN, EAB,PAA and FA and finally approved by all authors.

## REFERENCES

1. Meissner, W. et al. (2018) ‘Management of acute pain in the postoperative settingL: the importance of quality indicators’, Current Medical Research and Opinion, 0(0), pp. 187–196. Available at: 10.1080/03007995.2017.1391081.

2. Bach, A. M., Forman, A., & Seibaek, L. (2018). Postoperative Pain Management: A Bedside Perspective. Pain Management Nursing, 19(6), 608–618. 10.1016/j.pmn.2018.05.005

3. Rn, H. W. et al. (2017) ‘Postoperative pain experiences in Chinese adult patients after thoracotomy and video-assisted thoracic surgery’, (February), pp. 2744–2754. doi: 10.1111/jocn.13789.

4. Chou, R., Gordon, D.B., de Leon-Casasola, O.A., Rosenberg, J.M., Bickler, S., Brennan, T., Carter, T., Cassidy, C.L., Chittenden, E.H., Degenhardt, E. and Griffith, S., 2016. Management of Postoperative Pain: a clinical practice guideline from the American pain society, the American Society of Regional Anesthesia and Pain Medicine, and the American Society of Anesthesiologists’ committee on regional anesthesia, executive committee, and administrative council. The journal of pain, 17(2), pp.131–157.

5. Meissner, W., Coluzzi, F., Fletcher, D., Huygen, F., Morlion, B., Neugebauer, E., Pérez, A. M., Meissner, W., Coluzzi, F., Fletcher, D., Huygen, F., Neugebauer, E., Pérez, A. M., Pergolizzi, J., & Meissner, W. (2015). Improving the management of post-operative acute painL: priorities for change Commentary Improving the management of post-operative acute painL: priorities for change. 7995. 10.1185/03007995.2015.1092122

6. Málek, J., Ševčík, P., Bejšovec, D., Gabrhelík, T., Hnilicová, M., Křikava, I. and Mixa, V., 2017. Postoperative pain management. Prague, Czech Republic: Mladá fronta, pp.102-11.dicators. Current Medical Research and Opinion, 0(0), 187–196.

7. Liu, Q.-R., Ji, M.-H., Dai, Y.-C., Sun, X.-B., Zhou, C.-M., Qiu, X.-D., & Yang, J.-J. (2021).Predictors of Acute Postsurgical Pain following Gastrointestinal Surgery: A Prospective Cohort Study. Pain Research and Management, 2021, 1–8. 10.1155/2021/6668152

8. Rawal, N. (2016). Current issues in postoperative pain management. 160–171. 10.1097/EJA.0000000000000366

9. Mishchuk, V. R. (2017). Postoperative Pain in Children (Literature Review). Emergency Medicine, 0(8.79), 140–145. 10.22141/2224-0586.8.79.2016.90390

10. Mwashambwa, M. Y., Yongolo, I. M., Kapalata, S. N., & Meremo, A. J. (2018). Post-operative pain prevalence, predictors, management practices and satisfaction among operated cases at a regional referral hospital in Dar es Salaam, Tanzania. Tanzania Journal of Health Research, 20(2), 1–8. 10.4314/thrb.v20i2.10

11. Sitorus, F.E. et al. (2020) ‘The Problem Strategy and Emotion Focus Coping with Pain Intensity in Post Major Surgery’, (Ichimat 2019), pp. 521–527. Available at: 10.5220/0009974205210527.

12. Stallman, H. M. (2020). Health theory of coping. Australian Psychologist, 55(4), 295–306. 10.1111/ap.12465

13. Hess, E. P., Grudzen, C. R., Thomson, R., Raja, A. S., & Carpenter, C. R. (2015). Shared Decision-making in the Emergency. 856–864. 10.1111/acem.12703

14. Hour, O. C. (2016) ‘Pain Assessment and Management’.

15. Ninnoni, J.P.K. (2019) ‘Assessment and Management of Postoperative Pain among Nurses at a Resource-Constraint Teaching Hospital in Ghana’, 2019.

16. Aziato, L. and Adejumo, O. (2015) ‘An Ethnographic Exploration of Postoperative Pain Experiences Among Ghanaian Surgical Patients’, Journal of Transcultural Nursing, 26(3), pp. 301–307. doi: 10.1177/1043659614526246.

17. Menlah, A. et al. (2018) ‘Knowledge, Attitudes, and Practices of Postoperative Pain Management by Nurses in Selected District Hospitals in Ghana’, SAGE Open Nursing, 4, pp. 1–11. Available at: 10.1177/2377960818790383.

18. Kyei, E. F., Amponsah, A. K., & Boateng, E. A. (2019). “ I Experience Very Sharp Pain but It ’ s On and Off ”: A Phenomenological Study of Postoperative Pain Experiences of Patients in Ghana. March.

19. Neubauer, B. E., Witkop, C. T., & Varpio, L. (2019). How phenomenology can help us learn from the experiences of others. 90–97. 10.1007/s40037-019-0509-2

20. Rodriguez, A., & Smith, J. (2018). Phenomenology as a healthcare research method. 21(4), 96–98.

21. Prescott, F.J., 2011. Validating a long qualitative interview schedule. WoPaLP, 5, pp.16–38.

22. Vaismoradi, M. et al. (2016) ‘Theme development in qualitative content analysis and thematic analysis’, Journal of Nursing Education and Practice, 6(5). doi: 10.5430/jnep.v6n5p100.

23. Braun, V. and Clarke, V. (2012) ‘Thematic analysis’, 2. Available at: 10.1037/13620-004.

24. Creswell, J., 2009. Research design: Qualitative, quantitative, and mixed methods approaches 3rd Edition CA: Sage.

25. Guba, E.G. and Lincoln, Y.S., 1989. Fourth generation evaluation. Sage.

26. Bayman, E.O., Parekh, K.R., Keech, J., Larson, N., Vander Weg, M. and Brennan, T.J., 2019. Preoperative patient expectations of postoperative pain are associated with moderate to severe acute pain after VATS. Pain medicine, 20(3), pp.543–554.

27. Sipilä, R.M. et al. (2017) ‘Does expecting more pain make it more intense? Factors associated with the first week pain trajectories after breast cancer surgery’, Pain, 158(5), pp. 922–930. Available at: 10.1097/j.pain.0000000000000859.

28. Chatchumni, M. et al. (2016) ‘Thai Nurses’ experiences of post-operative pain assessment and its’ influence on pain management decisions’, BMC Nursing, 15(1), pp. 1–8. doi: 10.1186/s12912-016-0136-8.

29. Montgomery, G. H., Schnur, J. B., Erblich, J., Diefenbach, M. A., & Bovbjerg, D. H. (2010). Presurgery Psychological Factors Predict Pain, Nausea, and Fatigue One Week After Breast Cancer Surgery. Journal of Pain and Symptom Management, 39(6), 1043–1052. 10.1016/j.jpainsymman.2009.11.318

30. Ju, J. et al. (2018) ‘Factors affecting postoperative pain and delay in discharge from the post-anaesthesia care unitL: A descriptive correlational study’. doi: 10.1177/2010105817738794.

31. Gan, T.J., Habib, A.S., Miller, T.E., White, W. and Apfelbaum, J.L., 2014. Incidence, patient satisfaction, and perceptions of post-surgical pain: results from a US national survey. Current medical research and opinion, 30(1), pp.149–160.

32. Gamez, B. H., & Habib, A. S. (2018). Predicting severity of acute pain after cesarean delivery: A narrative review. Anesthesia and Analgesia, 126(5), 1606–1614. 10.1213/ANE.0000000000002658

33. Poudyal, S., Sharma, K., Subba, H.K. and Thapa, S., 2018. Pain experiences among postoperative patients admitted in surgical ward of teaching hospital, Chitwan. Asian Journal of Multidisciplinary Studies, 6(8), pp.1–7.

34. Subramanian, P., Ramasamy, S., Ng, K. H., Chinna, K., & Rosli, R. (2016). Pain experience and satisfaction with postoperative pain control among surgical patients. International Journal of Nursing Practice, 22(3), 232–238. 10.1111/ijn.12363

35. Eshete, M. T., Baeumler, P. I., Siebeck, M., Tesfaye, M., Haileamlak, A., Michael, G. G., Ayele, Y., & Irnich, D. (2019). Quality of postoperative pain management in Ethiopia: A prospective longitudinal study. PLoS ONE, 14(5), 1–22. 10.1371/journal.pone.0215563

36. Thamilselvam, P., & Pandurangan, T. (2017). Postoperative Pain Management. 1–3. 10.31031/DAPM.2017.01.000501

37. Widar, M. and Ek, A. (2004) ‘Coping with Long-Term Pain After a Stroke’, 27(3), pp. 215–225. doi: 10.1016/j.jpainsymman.2003.07.006.

38. Tabriz, E. R., Mohammadi, R., Roshandel, G. R., Talebi, R., & Khorshidi, M. (2018). Pain coping strategies and their relationship with unpleasant emotions (Anxiety, stress, and depression) and religious coping in cancer patients. Middle East Journal of Cancer, 9(3), 208–216. 10.30476/mejc.2018.42125

39. Dijkstra, M. and Homan, A.C., 2016. Engaging in rather than disengaging from stress: Effective coping and perceived control. Frontiers in psychology, 7, p.1415.

40. Pragholapati, A. (2020). Coping Strategies for Someone Divorced. 1985, 1–6. 10.31234/osf.io/w4zdb

41. Johnson, D. et al. (2018) ‘The impact of different coping strategies on acute and subacute post-mastectomy pain’, The Journal of Pain, 19(3), p. S76. Available at: 10.1016/j.jpain.2017.12.175

42. Shoqirat, N., Mahasneh, D., Singh, C. and Hadid, L. A. (2019). Do surgical patients’ characteristics and behaviours affect nurses’ pain management decisions? A qualitative inquiry. International Journal of Nursing Practice, 25(6), pp. 1–8. doi: 10.1111/ijn.12779.

43. Kaptain, K., Bregnballe, V. and Dreyer, P., 2017. Patient participation in postoperative pain assessment after spine surgery in a recovery unit. Journal of Clinical Nursing, 26(19-20), pp.2986–2994.

